# Impaired Skeletal Muscle Glucose Handling in Patients with Advanced Stage Lung Cancer Is Improved by Exercise Training Despite Blunted Molecular Plasticity

**DOI:** 10.64898/2026.11.19.26368511

**Authors:** Christian S. Carl, Anna M. H. Madsen, Jonas Sørensen, Nanna J. Hahn, Nicoline R. Andersen, Cecilie B. Lindqvist, Asker Damgaard, Morten Suhr, Mbulelo Karl Mfeketo, Marcus A. B. Hjorth, Sanne E. Høeg, Christian Lillelund, Morten Quist, Erik A. Richter, Lykke Sylow

## Abstract

**Background:** Patients with lung cancer often exhibit impaired glycemic control and insulin resistance, which are associated with poor clinical outcomes and increased mortality. However, skeletal muscle insulin sensitivity and the potential of exercise training to improve muscle glucose handling remain undetermined in these patients.

**Methods:** Ten patients with advanced stage lung cancer and 12 matched controls completed five weeks of one-legged knee-extension exercise training. Whole-body and leg insulin sensitivity were assessed by a hyperinsulinemic-euglycemic clamp with femoral arterio-venous catheterization and blood flow measurements. Muscle biopsies from both legs, obtained before and during insulin stimulation, were analyzed for glycogen, protein abundance, and insulin signaling.

**Results:** Compared with controls, patients had lower whole-body insulin sensitivity (-24%, p=0.030), leg glucose extraction (-37%, p=0.030), and muscle glycogen (-15%, p=0.045). Training increased leg glucose uptake (26%, p<0.001), blood flow (17%, p=0.005), and glycogen (32%, p<0.001) across both groups. Protein abundance did not differ between groups in the non-trained leg. Training increased GLUT4, Akt2, GSK3β, and AMPKα2 across both groups, whereas HKII and TBC1D4 increased only in controls, resulting in a 61% smaller overall molecular training response in patients (composite protein score, p=0.004). Training-induced changes in leg glucose uptake correlated with changes in GLUT4 levels (r=0.46, p=0.042). Insulin-stimulated phosphorylation of Akt^Ser473^, GSK3β^Ser9^, and TBC1D4^Thr642^ did not differ between groups.

**Conclusions:** Advanced stage lung cancer is associated with impaired whole-body and skeletal muscle glucose handling. Exercise training was effective in increasing muscle glucose uptake in these patients, although blunted molecular adaptations may limit the full metabolic benefit.

## Introduction

Lung cancer, the leading cause of cancer-related mortality worldwide, is associated with impaired glycemic control and an increased risk of type 2 diabetes (T2D), both of which are linked to poorer survival and higher recurrence rates (1–5). Although insulin resistance has been reported in patients with lung cancer (6), the site of impaired glucose handling and underlying mechanisms remain poorly understood, limiting the development of targeted interventions to improve metabolic control.

Skeletal muscle is responsible for the majority of insulin-stimulated glucose disposal in humans (7), yet skeletal muscle-specific insulin sensitivity has not been directly examined in patients with lung cancer. Addressing this gap is important because insulin resistance may promote compensatory hyperinsulinemia, a proposed driver of cancer development (8,9). While recent studies have described systemic and skeletal muscle alterations associated with early and advanced stage lung cancer (10,11), their contribution to insulin action remains unclear.

Exercise training is recommended guideline and a cornerstone treatment for alleviating insulin resistance in obesity, T2D, and other metabolic disorders (12,13), but its effects on insulin sensitivity in cancer have not been established. Therefore, we assessed whole-body and skeletal muscle insulin sensitivity in patients with advanced stage lung cancer and matched controls following five-weeks of one-legged exercise training. Insulin sensitivity was assessed using the gold-standard hyperinsulinemic-euglycemic clamp combined with femoral arteriovenous (a-v) balance and femoral blood flow measurements in both legs, enabling a direct comparison of the trained and non-trained leg. Muscle biopsies from both legs were analyzed for key proteins in glucose metabolism and insulin signaling and together, these findings indicate impaired skeletal muscle glucose handling in patients with advanced stage lung cancer.

### Patients with advanced stage lung cancer exhibit impaired skeletal muscle glucose handling, but training-induced improvements were preserved

In this controlled exercise training study, we compared metabolic and molecular responses in 10 patients with advanced stage lung cancer and 12 matched control participants without cancer. Groups were matched for sex (50% female), age (65.6 ± 6.5 years), body mass index (24.9 ± 3.5 kg·m^−2^), and cardiorespiratory fitness (30.2 ± 6.1 mL·min^−1^·kg^−1^) (Table S1). Pre-training peak workload did not differ between groups, and both groups completed an average of 15 training sessions at the same relative exercise intensities (Table S2).

To assess whole-body and skeletal muscle insulin sensitivity, participants underwent a hyperinsulinemic-euglycemic clamp combined with arterio-venous femoral catheterization and femoral artery blood flow measurements in the trained (T) and non-trained (NT) leg (Fig. 1A). Euglycemia was maintained throughout the clamp, with the glucose infusion rate (GIR) reaching steady state during the final 40 minutes of insulin stimulation (Fig. 1B).

**Figure 1.**
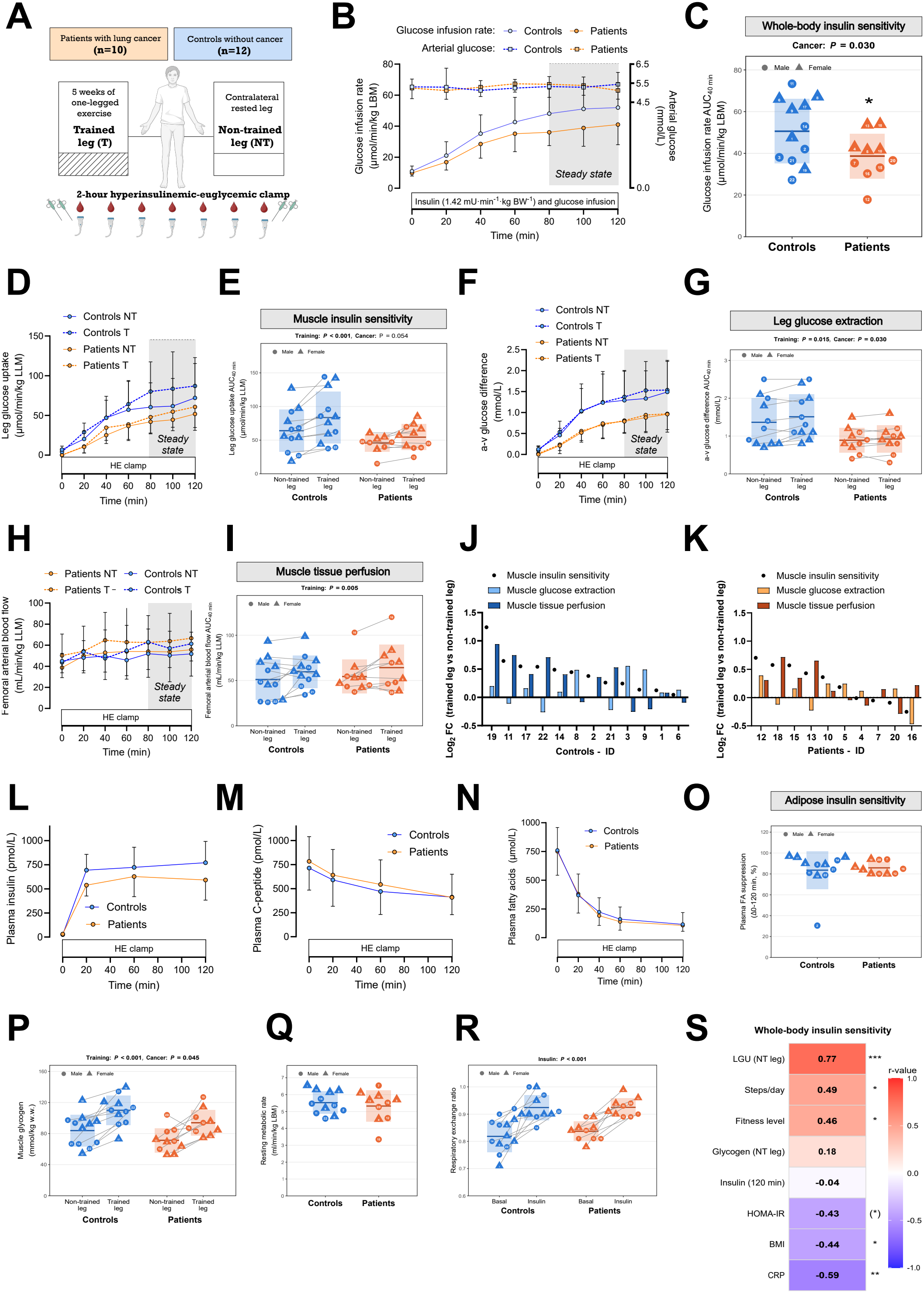
Patients with advanced stage lung cancer exhibit impaired skeletal muscle glucose handling, but training-induced improvements were preserved. **(A)** Study design. Ten patients with advanced stage lung cancer and 12 matched controls completed a five-week one-legged exercise training intervention. Whole-body and skeletal muscle insulin sensitivity were assessed by a 2-hour hyperinsulinemic-euglycemic (HE) clamp combined with femoral arterio-venous catheterization and femoral arterial blood flow measurements in the trained (T) and non-trained (NT) leg. **(B)** Glucose infusion rate and arterial plasma glucose concentrations during the HE clamp. **(C)** Whole-body insulin sensitivity assessed as glucose infusion rate (GIR) during steady-state insulin stimulation (80–120 min). **(D)** Time course of leg glucose uptake during the HE clamp and **(E)** muscle insulin sensitivity assessed as leg glucose uptake expressed as area under the curve during steady state. **(F)** Time course of glucose extraction (arterio-venous (a-v) glucose difference) and **(G)** glucose extraction expressed as area under the curve during steady state. **(H)** Time course of femoral arterial blood flow and **(I)** femoral arterial blood flow expressed as area under the curve during steady state. **(J)** Individual training-induced log2 fold-changes (trained versus non-trained leg) in muscle insulin sensitivity, glucose extraction, and arterial blood flow in controls and **(K)** patients, ordered by the change in muscle insulin sensitivity. Numbers on the x-axis are participant IDs. **(L)** Plasma insulin and **(M)** plasma C-peptide concentrations during the HE clamp. **(N)** Plasma fatty acid (FA) concentrations during the HE clamp and **(O)** adipose tissue insulin sensitivity assessed as the suppression of plasma FA from 0 to 120 min (%). **(P)** Muscle glycogen content in non-trained and trained legs. **(Q)** Resting metabolic rate and **(R)** respiratory exchange ratio (RER) at baseline and during insulin stimulation. **(S)** Correlations between whole-body insulin sensitivity and selected clinical and metabolic variables. Partial correlation coefficients adjusted for sex are shown within the heatmap (Pearson; Spearman for CRP, as most values were at the detection limit); n = 22 (fitness level, n = 21). Data are presented as mean ± SD. Individual data points are shown with participant ID; circles indicate males and triangles females. Time courses (B, D, F, H, L–N) and individual fold changes (J–K) are descriptive. Group differences in C, O and Q were assessed using linear models adjusted for sex. Main effects of group and training (E, G, I, P) or insulin (R) and their interaction were assessed using linear mixed-effects models adjusted for sex, with participant as a random effect. (^*^) p<0.10, ^*^ p<0.05, ^**^ p<0.01, ^***^ p<0.001. AUC, area under the curve; BW, body weight; CRP, C-reactive protein; FA, fatty acids; GIR, glucose infusion rate; HOMA-IR, homeostatic model assessment of insulin resistance; LBM, lean body mass; LGU, leg glucose uptake; LLM, lean leg mass; NT, non-trained; T, trained. Illustration in (A) was created using BioRender.

Whole-body insulin sensitivity, quantified as GIR at steady state, was 24% lower (p=0.030) in patients with lung cancer compared with controls (Fig. 1C). Similarly, skeletal muscle insulin sensitivity, assessed as leg glucose uptake tended to be 32% lower in patients (p=0.054; Fig. 1D–E). The estimated group difference was -23.7 µmol/min/kg LLM (95% CI: -48.2 to 0.8), compatible with a physiologically meaningful reduction but with considerable inter-individual variability. Exercise training increased leg glucose uptake with 26% (p<0.001; Fig. 1D-E) across both groups. These data support previous findings of whole-body insulin resistance in patients with lung cancer (6) even when carefully controlling for parameters known to influence insulin sensitivity like sex, age, BMI, fitness level, and standardizing diet before testing (see Methods). Furthermore, we extend previous observations by demonstrating a strong tendency towards skeletal muscle insulin resistance in patients with lung cancer, which could be counteracted by exercise training.

Using the arterio-venous balance technique, leg glucose uptake is composed of glucose extraction (arterio-venous glucose difference) and femoral arterial blood flow. Patients exhibited 37% lower glucose extraction compared with controls (p=0.030), with exercise training increasing glucose extraction by 8% across both groups (p=0.015, Fig. 1F-G). Exercise training also increased femoral arterial blood flow by 17%, compared with the non-trained legs across both groups (p=0.005, Fig. 1H–I). Individual responses in the two groups revealed marked heterogeneity in the training adaptation of these two components and no clear differences between patients and controls (Fig. 1J-K).

In patients with T2D, leg glucose extraction, but not leg blood flow, was impaired at baseline and during insulin stimulation, resulting in lower leg glucose clearance compared with controls. One-legged high-intensity interval training increased insulin-stimulated leg glucose clearance by ∼30% in both groups, mainly through increased leg blood flow with maintained glucose extraction (14). Furthermore, in patients with mitochondrial dysfunction, both leg glucose extraction and leg blood flow were lower during insulin stimulation in the non-trained state then in matched controls, but only leg blood flow, and not glucose extraction, tended to increase following exercise training (15). Together, these findings suggest that impaired glucose extraction may be a common feature of skeletal muscle insulin resistance across diverse disease states, and that exercise training primarily improves leg blood flow rather than glucose extraction.

Despite receiving comparable relative insulin infusion rates, patients tended to exhibit lower steady-state arterial plasma insulin concentrations (p=0.083 at 120 minutes time point; Fig. 1L), a finding that has been observed previously (16). As endogenous insulin secretion during the clamp, measured by plasma C-peptide levels, was similar in patients and controls (Fig. 1M), which may indicate increased insulin clearance in the patients. Adipose tissue insulin sensitivity, measured as insulin’s ability to suppress fatty acid mobilization, was preserved in patients (Fig. 1N-O).

In the basal state, glycogen concentration was 15% lower (p=0.045) in patients compared to controls but exercise training increased glycogen concentration with 32% (p<0.001) across both groups (Fig. 1P). Resting metabolic rate and respiratory exchange ratio (RER) measured by indirect calorimetry were similar between groups and RER increased to the same extent with insulin stimulation across both groups (Fig. 1Q-R), suggesting that whole-body carbohydrate oxidation was not compromised in patients in either the basal or the insulin-stimulated state.

Whole-body insulin sensitivity correlated strongly with glucose uptake in the non-trained leg (r=0.77, p<0.001) and moderately with daily physical activity (steps/day; r=0.49, p=0.025) and fitness level (r=0.46, p=0.041), and was inversely associated with C-reactive protein (CRP) (r=-0.59, p=0.005) and body mass index (BMI) (r=-0.44, p=0.047) and tended to be inversely associated with homeostatic model assessment of insulin resistance (HOMA-IR) (r=-0.43, p=0.054) (Fig. 1S). Patients took fewer daily steps and had higher CRP than controls (Table S1). Low habitual physical activity has previously been reported in patients with lung cancer (17), and CRP, a marker of low-grade inflammation, is inversely associated with insulin sensitivity (18), suggesting that both may have contributed to the lower insulin sensitivity in patients. Notably, end-clamp insulin concentrations were not associated with insulin sensitivity (Fig. 1S), suggesting that group differences in insulin action were independent of individual differences in circulating insulin concentrations during the clamp.

### Exercise training increases skeletal muscle protein abundance, but with blunted adaptations in patients

Exercise training enhances insulin sensitivity through upregulation of key proteins involved in skeletal muscle glucose uptake and intracellular glucose metabolism (19,20). Protein abundance of key metabolic proteins was not different between patients and controls in the non-trained leg. However, exercise training increased glucose transporter 4 (GLUT4; +34%, p<0.001), AKT serine/threonine kinase 2 (Akt2; +52%, p<0.001), glycogen synthase kinase 3 beta (GSK3β; +26%, p=0.008), and 5’-AMP-activated protein kinase α2 (AMPKα2; +36%, p<0.001) across both groups, whereas hexokinase II (HKII; +77%, p<0.001 in controls vs. +29%, p=0.49 in patients) and TBC1 domain family member 4 (TBC1D4; +54%, p<0.001 vs. +17%, p=0.30) only increased in the controls (Fig. 2A-F). An overall composite protein score showed a 61% smaller training response in patients than in controls (training × cancer interaction, p=0.004) compared with controls (Fig. 2G). These data indicate that protein abundance is not affected by advanced stage lung cancer in the non-trained state, but the exercise response seems blunted for at least some of the proteins involved in glucose metabolism. Resistance to molecular training adaptations has been observed in insulin-resistant individuals with T2D: two weeks of one-legged high-intensity interval training induced an attenuated myonuclear transcriptional response compared with healthy controls, particularly in genes regulating glycogen metabolism and glycolysis, despite a similar improvement in glucose clearance (21). However, whether this blunted training response was confined to the proteins measured here or reflects a broader, proteome-wide attenuation of the training response remains to be determined. Identifying the mechanisms underlying this blunted response may reveal novel therapeutic targets to restore exercise responsiveness and maximize the clinical benefit of exercise training in this patient population.

**Figure 2.**
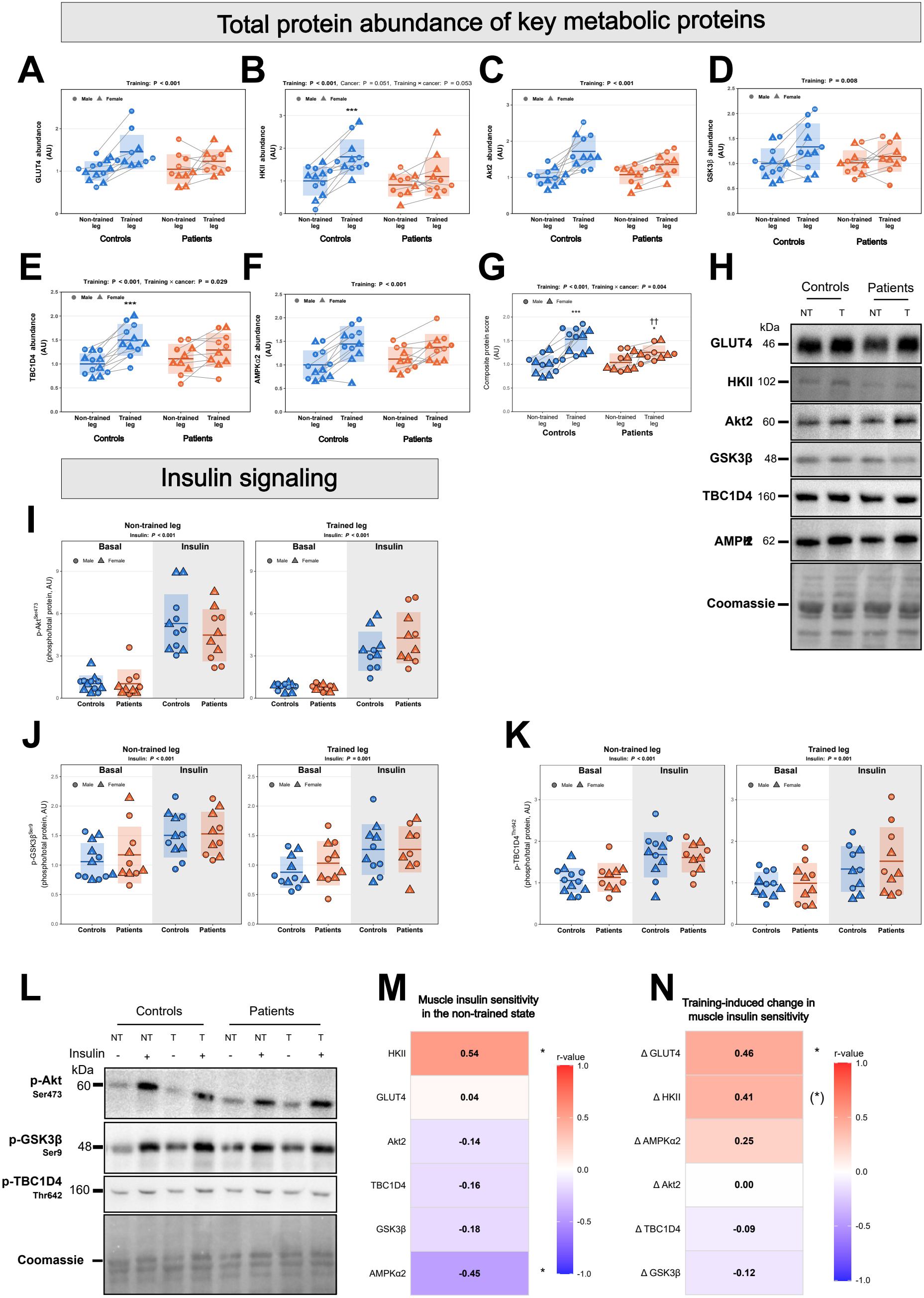
Exercise training increases skeletal muscle protein abundance, but with blunted adaptations in patients. Protein abundance of GLUT4 **(A)**, HKII **(B)**, Akt2 **(C)**, GSK3β **(D)**, TBC1D4 **(E)**, and AMPKα2 **(F)** in non-trained (NT) and trained (T) legs from controls and patients with advanced stage lung cancer. **(G)** Composite protein score, calculated for each participant and leg as the mean abundance of all six measured proteins. **(H)** Representative immunoblots for the proteins shown in panels A–F. Blot for GSK3β is cropped because insulin-stimulated samples were loaded together with basal samples **(I–K)**. Phosphorylation of Akt Ser473 **(I)**, GSK3β Ser9 **(J)**, and TBC1D4 Thr642 **(K)** relative to total protein under basal and insulin-stimulated conditions in the non-trained and trained leg of controls and patients. **(L)** Representative immunoblots for phosphorylated proteins shown in panels I–K. Coomassie staining was used as a loading control. **(M)** Correlations between skeletal muscle insulin sensitivity (leg glucose uptake during steady state) in the non-trained leg and protein abundance in the non-trained leg. **(N)** Correlations between the training-induced change in skeletal muscle insulin sensitivity and training-induced changes in protein abundance (Δ, trained - non-trained leg). Partial Pearson correlation coefficients (r) adjusted for sex are shown within each heatmap. Data are presented as individual values with mean ± SD; circles indicate males and triangles females. In A–G, main effects of group and training and their interaction were assessed using linear mixed-effects models adjusted for sex, with participant as a random effect. In I– K, main effects of group and insulin and their interaction were assessed separately for each leg using the same type of model. When the interaction had p<0.05 (p=0.053 in B), Sidak-adjusted post hoc comparisons were conducted. ^*^ p<0.05, ^***^ p<0.001 trained vs non-trained leg within the group; †† p<0.01 patients vs controls in the same leg (G). (^*^) p<0.10, ^*^ p<0.05 (M–N). Protein abundance and phosphorylation were quantified and expressed in arbitrary units (AU) relative to the non-trained leg in the controls (basal for phosphorylation). AUC, area under the curve; T, trained leg; NT, non-trained leg; GLUT4, glucose transporter 4; HKII, hexokinase II; Akt2, AKT serine/threonine kinase 2; GSK3β, glycogen synthase kinase 3β; TBC1D4, TBC1 domain family member 4; AMPKα2, 5′-AMP-activated protein kinase α2.

Insulin stimulation increased phosphorylation of Akt Ser^473^, GSK3β Ser^9^, and TBC1D4 Thr^642^ in both controls and patients, with no difference in the response between the two groups (Fig. 2I-K). Thus, insulin signaling at the measured sites in skeletal muscles was not affected by cancer in this patient cohort. In contrast, pre-clinical studies have shown that insulin-induced TBC1D4 and GSK3 phosphorylation was abolished in soleus muscles from tumor-bearing mice along with abrogated insulin-stimulated glucose uptake (22). Whether this discrepancy reflects differences in cancer type, species, or muscle fiber type remains undefined.

Muscle insulin sensitivity in the non-trained state was positively associated with HKII abundance (r=0.54, p=0.012) and negatively associated with AMPKα2 abundance (r=-0.45, p=0.041) (Fig. 2M; Fig. S1A-B). Patients with non-small cell lung cancer have been reported to have a higher muscle content of several AMPK subunits, including α2 (23). In tumor-bearing mice, loss of muscle AMPK accelerated cancer-associated insulin resistance (23), indicating that muscle AMPK counteracts cancer-induced metabolic disturbances. The negative association between AMPKα2 abundance and insulin sensitivity observed in the present study may therefore reflect a compensatory upregulation of AMPK in the most insulin-resistant participants, which is insufficient to fully preserve insulin sensitivity. The training-induced changes in muscle insulin sensitivity were associated with increased GLUT4 abundance (r=0.46, p=0.042) and tended to be associated with increased HKII abundance (r=0.41, p=0.072) (Fig. 2N; Fig. S1C-D). That training-induced improvements in insulin sensitivity correlate with increased GLUT4 abundance has previously been reported in healthy young men (24), but we show that this correlation is still evident in an older population with and without advanced stage lung cancer. Furthermore, HKII has also been discussed as a rate-limiting step for insulin-stimulated glucose uptake (25), and we show that HKII abundance correlates with insulin-stimulated glucose uptake in the non-trained state and that it tended to correlate with training-induced adaptations, linking protein adaptations to increased insulin-stimulated glucose uptake following exercise training.

Collectively, these findings demonstrate that patients exhibit blunted training-induced molecular adaptations of proteins involved in muscle glucose metabolism, but insulin signaling seems to be unaffected by advanced stage lung cancer.

### Strength and Limitations

A major strength of this study is that it is the first to measure skeletal muscle glucose uptake using gold standard technique, and the first to study the effects of supervised one-legged exercise training on muscle insulin sensitivity in patients with advanced stage lung cancer. The one-legged training model allowed direct within-subject comparisons between trained and non-trained muscles, minimizing inter-individual variability, strengthening statistical power, and reducing participant burden by requiring only a single hyperinsulinemic-euglycemic clamp experiment.

Recruitment was challenging due to the demanding intervention and invasive procedures, and selection bias cannot be excluded, as participants may have represented a healthier and more resourceful subgroup of patients. However, baseline characteristics were comparable to those of patients recruited from the same clinic to a less invasive study (10), suggesting limited selection bias. Further limitations include the small sample size, heterogeneity in tumor type (non-small cell lung cancer (NSCLC), small cell lung cancer (SCLC) and mesothelioma) and ongoing anti-cancer treatment, and the exploratory correlation analyses, which were not adjusted for multiple comparisons.

## Conclusion

The present investigation provides evidence that skeletal muscle glucose handling is impaired in patients with advanced stage lung cancer. Furthermore, exercise training improved muscle insulin action, supporting its potential as a therapeutic strategy to counteract cancer-associated insulin resistance. These findings support integrating supervised exercise training into the care of patients with advanced stage lung cancer, while the blunted molecular training response warrants investigation of its underlying mechanisms to maximize the metabolic benefit of exercise in this population.

## Supporting information

Supplemental table 1, 2, and 3, and method section

## Data Availability

All data produced in the present study are available upon reasonable request to the authors

## Acknowledgements

We thank the nurses and physicians at the Department of Oncology, Copenhagen University Hospital Rigshospitalet, for their invaluable support during participant recruitment and inclusion, and the staff at the Department of Clinical Biochemistry, Rigshospitalet, Copenhagen, Denmark for blood analyses. We thank Irene Bech Nielsen from the Department of Nutrition, Exercise, and Sports for analyzing insulin and fatty acid concentrations, and Simon Bech Petersen from the Department of Biomedical Sciences for analyzing c-peptide. We also thank the students who assisted with the training sessions and helped create a safe and supportive environment for the participants.

Finally, we are deeply grateful to all study participants for their time and commitment. We especially acknowledge the patients who took part in this study. Through their willingness to contribute despite the challenges associated with their disease, they have helped generate knowledge that may improve the care and treatment of future patients with cancer.

L.S. was supported by the Independent Research Fund Denmark (0169-00060B, 3146-00040B, 0169-00013B) and the Danish Cancer Society (R302-A17605). L.S. was supported by Sejer Persson and Lis Klüwer Perssons Legat. The funds were not involved in the study or manuscript in any way.

## Author contributions

Conceptualization: L.S. and J.S. Data curation: C.S.C., A.M. H.M., C.B.L., A.D., M.S., M.K.M., and M.A.B.H. Formal analysis: C.S.C., A.M. H.M., C.B.L., A.D., and M.S. Funding acquisition: L.S. Investigation: C.S.C., A.M.H.M., N.J.H., C.B.L., A.D., M.S., M.K.M., M.A.B.H., N.R.A., and E.A.R. Methodology: L.S., J.S., C.S.C., A.M.H.M., N.R.A., and E.A.R. Project administration: C.S.C., A.M.H.M., and L.S. Resources: L.S., J.S., C.S.C., A.M.H.M., S.E.H., C.L., and M.Q. Software: C.S.C., A.M.H.M., C.B.L., A.D., and M.S. Supervision: L.S. Validation: C.S.C. Visualization: C.S.C., C.B.L., A.D., and M.S. Writing – original draft: C.S.C., L.S., and A.M.H.M. Writing – review and editing: C.S.C., A.M.H.M., J.S., N.J.H., N.R.A., C.B.L., A.D., M.S., M.K.M., M.A.B.H., S.E.H., C.L., M.Q., E.A.R., and L.S.

## Declaration of generative AI and AI-assisted technologies in the manuscript preparation process

During the preparation of this work the authors used Copilot (Microsoft) and Claude (Anthropic) in order to improve the language, grammar, and sentence structure of the manuscript. After using this tool/service, the authors reviewed and edited the content as needed and take full responsibility for the content of the published article.

## Reference list

1. Luo J, Chen YJ, Chang LJ. Fasting blood glucose level and prognosis in non-small cell lung cancer (NSCLC) patients. Lung Cancer. 2012 May;76(2):242–7. doi:10.1016/j.lungcan.2011.10.019 PubMed PMID: 22112292.

2. Bergamino M, Rullan AJ, Saigí M, Peiró I, Montanya E, Palmero R, et al. Fasting plasma glucose is an independent predictor of survival in patients with locally advanced non-small cell lung cancer treated with concurrent chemoradiotherapy. BMC Cancer. BioMed Central Ltd.; 2019. doi:10.1186/s12885-019-5370-5 PubMed PMID: 30791870.

3. Hwangbo Y, Kang D, Kang M, Kim S, Lee EK, Kim YA, et al. Incidence of Diabetes After Cancer Development: A Korean National Cohort Study. JAMA Oncol. 2018 Aug 1;4(8):1099–105. doi:10.1001/jamaoncol.2018.1684 PubMed PMID: 29879271.

4. Sylow L, Grand MK, von Heymann A, Persson F, Siersma V, Kriegbaum M, et al. Incidence of New-Onset Type 2 Diabetes After Cancer: A Danish Cohort Study. Diabetes Care. 2022 Jun 2;45(6):e105–6. doi:10.2337/dc22-0232 PubMed PMID: 35653597.

5. Varlotto J, Medford-Davis LN, Recht A, Flickinger J, Schaefer E, Shelkey J, et al. Confirmation of the role of diabetes in the local recurrence of surgically resected non-small cell lung cancer. Lung Cancer. 2012 Mar;75(3):381–90. doi:10.1016/j.lungcan.2011.07.019 PubMed PMID: 21864933.

6. Màrmol JM, Carlsson M, Raun SH, Grand MK, Sørensen J, Lang Lehrskov L, et al. Insulin resistance in patients with cancer: a systematic review and meta-analysis. Acta Oncol (Madr). 2023;62(4). doi:10.1080/0284186X.2023.2197124

7. Honka MJ, Latva-Rasku A, Bucci M, Virtanen KA, Hannukainen JC, Kalliokoski KK, et al. Insulin-stimulated glucose uptake in skeletal muscle, adipose tissue and liver: A positron emission tomography study. Eur J Endocrinol. 2018 May 1;178(5):523–31. doi:10.1530/EJE-17-0882 PubMed PMID: 29535167.

8. Zhong W, Mao Y. Daily Insulin Dose and Cancer Risk Among Patients With Type 1 Diabetes. JAMA Oncol. 2022;8(9). doi:10.1001/jamaoncol.2022.2960

9. Zhang AMY, Magrill J, de Winter TJJ, Hu X, Skovsø S, Schaeffer DF, et al. Endogenous Hyperinsulinemia Contributes to Pancreatic Cancer Development. Cell Metabolism. 2019. doi:10.1016/j.cmet.2019.07.003

10. Sørensen J, Voldstedlund CT, Ogueboule ZKJ, Modvig J, Knøs C, Hammershøi A, et al. Plasma proteomics identifies an IL-6–associated SAA axis linked to muscle wasting in patients with cancer cachexia. bioRxiv. 2026 Jan 1;2026.06.11.730788. doi:10.64898/2026.06.11.730788

11. Al-Sawaf O, Weiss J, Skrzypski M, Lam JM, Karasaki T, Zambrana F, et al. Body composition and lung cancer-associated cachexia in TRACERx. Nat Med. 2023 Apr 1;29(4):846–58. doi:10.1038/s41591-023-02232-8 PubMed PMID: 37045997.

12. Kanaley JA, Colberg SR, Corcoran MH, Malin SK, Rodriguez NR, Crespo CJ, et al. Exercise/Physical Activity in Individuals with Type 2 Diabetes: A Consensus Statement from the American College of Sports Medicine. Med Sci Sports Exerc. 2022 Feb 1;54(2):353–68. doi:10.1249/MSS.0000000000002800 PubMed PMID: 35029593.

13. Vainshtein A, Blaauw B, De Bock K, Munoz-Canoves P, Olson EN, Ottenheijm CAC, et al. The hallmarks of skeletal muscle health. Nature Metabolism. Nature Research; 2026. doi:10.1038/s42255-026-01595-9

14. Dela F, Ingersen A, Andersen NB, Nielsen MB, Petersen HHH, Hansen CN, et al. Effects of one-legged high-intensity interval training on insulin-mediated skeletal muscle glucose homeostasis in patients with type 2 diabetes. Acta Physiologica. 2019 Jun 1;226(2). doi:10.1111/apha.13245 PubMed PMID: 30585698.

15. Nielsen TL, Damgaard M, Andersen NR, Dunø M, Frederiksen AL, van Hall G, et al. An exercise-modifiable mitochondrial basis for skeletal muscle insulin resistance in humans. medRxiv. 2026 Jan 1;2025.12.17.25342274. doi:10.64898/2025.12.17.25342274

16. Winter A, MacAdams J, Chevalier S. Normal protein anabolic response to hyperaminoacidemia in insulin-resistant patients with lung cancer cachexia. Clinical Nutrition. 2012 Oct;31(5):765–73. doi:10.1016/j.clnu.2012.05.003 PubMed PMID: 22647419.

17. Granger CL, Parry SM, Denehy L. The self-reported Physical Activity Scale for the Elderly (PASE) is a valid and clinically applicable measure in lung cancer. Supportive Care in Cancer. 2015 Nov 30;23(11):3211–8. doi:10.1007/s00520-015-2707-8 PubMed PMID: 25808104.

18. Festa A, Ralph D’agostino;, Howard G, Mykkänen L, Tracy RP, Haffner SM. Chronic Subclinical Inflammation as Part of the Insulin Resistance Syndrome The Insulin Resistance Atherosclerosis Study (IRAS) [Internet]. 2000. Available from: http://www.circulationaha.org

19. Deshmukh AS, Steenberg DE, Hostrup M, Birk JB, Larsen JK, Santos A, et al. Deep muscle-proteomic analysis of freeze-dried human muscle biopsies reveals fiber type-specific adaptations to exercise training. Nat Commun. 2021;12(1). doi:10.1038/s41467-020-20556-8 PubMed PMID: 33436631.

20. Sylow L, Tokarz VL, Richter EA, Klip A. The many actions of insulin in skeletal muscle, the paramount tissue determining glycemia. Cell Metab. 2021 Apr 6;33(4):758–80. doi:10.1016/J.CMET.2021.03.020 PubMed PMID: 33826918.

21. Hansen M R Grothen JE, Karlsen A, Martinez JM, Sidiropoulos N, Helge JW, et al. The skeletal muscle response to high-intensity training assessed by single-nucleus RNA-sequencing is blunted in individuals with type 2 diabetes. J Physiol. 2025;603:3357–77. doi:10.1113/JP288368#support-information-section

22. Raun SH, Knudsen JR, Han X, Jensen TE, Sylow L. Cancer causes dysfunctional insulin signaling and glucose transport in a muscle-type-specific manner. FASEB Journal. 2022 Mar 1;36(3). doi:10.1096/fj.202101759R PubMed PMID: 35195922.

23. Raun SH, Ali MS, Han X, Henríquez-Olguín C, Pham TCP, Meneses-Valdés R, et al. Adenosine monophosphate-activated protein kinase is elevated in human cachectic muscle and prevents cancer-induced metabolic dysfunction in mice. J Cachexia Sarcopenia Muscle. 2023 Aug 1;14(4):1631–47. doi:10.1002/jcsm.13238

24. Dela F, Handberg A, Mikinest KJ, Galbo$ H. GLUT 4 AND INSULIN RECEPTOR BINDING AND KINASE ACTIVITY IN TRAINED HUMAN MUSCLE. Journal of Physiology. 1993.

25. Wasserman DH. Insulin, Muscle Glucose Uptake, and Hexokinase: Revisiting the Road Not Taken. Physiology. 2022 May 1;37(3):115–27. doi:10.1152/physiol.00034.2021 PubMed PMID: 34779282.

26. Björkman F, Ekblom-Bak E, Ekblom Ö, Ekblom B. Validity of the revised Ekblom Bak cycle ergometer test in adults. Eur J Appl Physiol. 2016;116(9). doi:10.1007/s00421-016-3412-0

27. Brzycki M. Strength Testing—Predicting a One-Rep Max from Reps-to-Fatigue. J Phys Educ Recreat Dance. 1993 Jan 1;64(1):88–90. doi:10.1080/07303084.1993.10606684

28. Fritzen AM, Lundsgaard AM, Jeppesen J, Christiansen MLB, Biensø R, Dyck JRB, et al. 5′-AMP activated protein kinase α2 controls substrate metabolism during post-exercise recovery via regulation of pyruvate dehydrogenase kinase 4. Journal of Physiology. 2015;593(21). doi:10.1113/JP270821

