## Supplemental figure 1 for "Impaired Skeletal Muscle Glucose Handling in Patients with Advanced Stage Lung Cancer Is Improved by Exercise Training Despite Blunted Molecular Plasticity"

### Figure S1. Correlations between skeletal muscle protein abundance and muscle insulin sensitivity

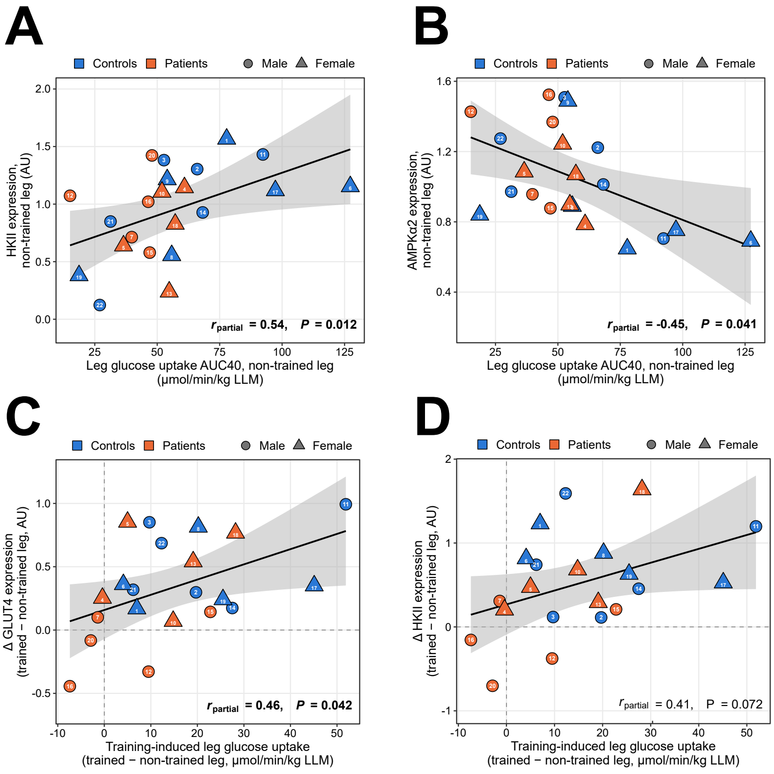
