## Supplemental table 1, 2, and 3, and method section for "Impaired Skeletal Muscle Glucose Handling in Patients with Advanced Stage Lung Cancer Is Improved by Exercise Training Despite Blunted Molecular Plasticity"

**Figure S1. Correlations between skeletal muscle protein abundance and muscle insulin sensitivity**

Correlations between insulin-stimulated leg glucose uptake during steady-state (AUC40) and HKII **(A)** and AMPKα2 **(B)** protein abundance in the non-trained leg. Correlations between training-induced changes (Δ, trained - non-trained leg) in insulin-stimulated leg glucose uptake (AUC40) and training-induced changes in GLUT4 **(C)** and HKII **(E)** protein abundance. Correlations were assessed using partial Pearson's correlation adjusted for sex (r_partial); n = 22 (A–B) and n = 21 (C–D). Data points represent individual participants with participant ID, with controls and patients indicated by color and males and females indicated by symbol shape. Solid lines represent linear regression fits with 95% confidence bands of the unadjusted data. AUC, area under the curve; GLUT4, glucose transporter 4; HKII, hexokinase II; AMPKα2, 5′-AMP-activated protein kinase α2; LLM, lean leg mass.

### Tables and legends

**Table S1 – Demographic and Clinical Characteristics of the Study Population at Baseline**

| **Characteristics** |  | **Units** | **Controls**  **(n=12)** | **Patients**  **(n=10)** | **p-value** |
| --- | --- | --- | --- | --- | --- |
| **Demographics** |  |  |  |  |  |
|  | Age | Years | 64.9 ± 6.4 | 66.4 ± 6.8 | 0.600 |
|  | Male/Female | n (%) | 6 / 6 (50/50) | 5 / 5 (50/50) |  |
| **Anthropometric measurements** |  |  |  |  |  |
|  | Weight | kg | 75.4 ± 16.1 | 74.2 ± 14.3 | 0.846 |
|  | BMI | kg · m^-2^ | 25.0 ± 4.0 | 24.8 ± 3.0 | 0.918 |
|  | Fat mass, DXA | % | 31.6 ± 8.1 | 33.1 ± 6.7 | 0.653 |
|  | Lean mass, DXA | % | 64.8 ± 7.9 | 63.5 ± 6.2 | 0.669 |
| **Physical activity level** |  |  |  |  |  |
|  | VO_2_-peak | l · min^-1^ | 2.4 ± 0.6 | 2.3 ± 0.7 | 0.743 |
|  | Fitness level | ml · min^-1^ · kg^-1^ | 31.0 ± 6.7 | 29.1 ± 5.4 | 0.513 |
|  | Daily physical activity | Steps | 10294 ± 4375 | 6885 ± 2851 | **0.047** |
| **Plasma parameters** |  |  |  |  |  |
|  | CRP | mg/L | 2.7 ± 5.3 | 48.3 ± 61.6 | **0.018** |
|  | Glucose (fasting) | mmol/L | 5.3 ± 0.4 | 5.2 ± 0.6 | 0.742 |
|  | Insulin (fasting) | pmol/L | 25.2 ± 9.4 | 30.6 ± 11.0 | 0.227 |
| **Calculated index** |  |  |  |  |  |
|  | HOMA-IR | AU | 1.0 ± 0.4 | 1.3 ± 0.5 | 0.266 |

Data are presented as mean ± SD. For CRP, eight out of 12 control participants had concentrations below the detection limit (<1 mg/L) and were assigned a value of 0.01 mg/L. Group differences were assessed using unpaired t-tests. Abbreviations: AU, arbitrary units; BMI, body mass index; CRP, C-reactive protein; DXA, dual-energy X-ray absorptiometry; HOMA-IR, homeostatic model assessment of insulin resistance; VO₂-peak, peak oxygen uptake.

**Table S2 – Training Characteristics in Patients with Advanced Stage Lung Cancer and Controls**

|  |  | **Units** | **Controls (n=12)** | **Patients (n=10)** | **p-value** |
| --- | --- | --- | --- | --- | --- |
| **Training** | |  |  |  |  |
|  | Training sessions | # | 15.0 ± 1.0 | 15.3 ± 1.3 | 0.546 |
| **One-legged leg extension (endurance)** | |  |  |  |  |
|  | Peak workload (PWL) | W | 28.3 ± 10.3 | 22.0 ± 10.9 | 0.176 |
|  | Average intensity | % of PWL | 94.1 ± 13.9 | 90.4 ± 6.5 | 0.446 |
| **Intensity levels (endurance)** | |  |  |  |  |
|  | ≤ 85 % of PWL | % of total work | 45.9 ± 14.8 | 49.0 ± 17.0 | 0.644 |
|  | 85-105 % of PWL | % of total work | 41.1 ± 14.6 | 42.0 ± 14.2 | 0.885 |
|  | ≥ 105 % PWL | % of total work | 13.0 ± 14.3 | 8.9 ± 3.9 | 0.392 |
| **One-legged leg extension (strength)** | |  |  |  |  |
|  | 10 Repetition Maximum (RM) | kg | 22.7 ± 10.0 | 19.3 ± 7.8 | 0.399 |
|  | Average intensity | % of 10 RM | 83.7 ± 10.2 | 84.4 ± 3.6 | 0.824 |
| **Intensity levels (strength)** | |  |  |  |  |
|  | ≤ 85 % of 10 RM | % of total work | 51.1 ± 27.5 | 55.8 ± 17.8 | 0.646 |
|  | 85-105 % of 10 RM | % of total work | 41.8 ± 24.9 | 36.7 ± 16.4 | 0.580 |
|  | ≥ 105 % of 10 RM | % of total work | 7.0 ± 4.5 | 7.5 ± 4.3 | 0.808 |

Data are presented as mean ± SD. Group differences were assessed using unpaired t-tests. Abbreviations: PWL, peak workload; RM, repetition maximum; W, watt.

**Table S3 – Overview of Antibodies and Dilutions**

| **Antibody** | **Company** | **Number #** | **Dilution** |
| --- | --- | --- | --- |
| GLUT4 | Thermo Fisher Scientific | PA1-1065 | 1:1000 in 2% skim milk |
| HKII | Cell Signaling Technology | #2867 | 1:1000 in 2% skim milk |
| Akt2 | Cell Signaling Technology | #3063s | 1:1000 in 2% skim milk |
| p-Akt Ser473 | Cell Signallng Technology | #9271 | 1:1000 in 2% skim milk |
| GSK3β | Bd Biosciences | #610202 | 1:1000 in 2% skim milk |
| p-GSK3β Ser9 | Cell Signaling Technology | #5558 | 1:1000 in 2% skim milk |
| TBC1D4 | Abcam | ab189890 | 1:2000 in 2% skim milk |
| p-TBC1D4 Thr642 | Cell Signaling Technology | #8881S | 1:1000 in 2% skim milk |
| AMPKα2 | Abcam | ab3760 | 1:1250 in 2% skim milk |

Abbreviations: GLUT4, glucose transporter 4; HKII, hexokinase II; Akt2, AKT serine/threonine kinase 2; GSK3β, glycogen synthase kinase 3β; TBC1D4, TBC1 domain family member 4; AMPKα2, 5′-AMP-activated protein kinase α2.

### Methods

#### Participants

Patients with advanced stage lung cancer were recruited from the Department of Oncology, Rigshospitalet, Copenhagen, Denmark. The study was approved by the Regional Ethics Committee of the Capital Region of Denmark (H-21035808), registered at ClinicalTrials.gov (NCT05307367), and conducted in accordance with the Declaration of Helsinki. Compliance with data protection regulations was ensured through a data processing agreement with the Capital Region of Denmark. All participants provided written informed consent prior to participation

Male and female patients with advanced stage (stage III-IV) lung cancer were included. The patient group comprised individuals diagnosed with non-small cell lung cancer (NSCLC, n=6), small cell lung cancer (SCLC, n=2), or malignant pleural mesothelioma (MPM, n=2). Eight of the ten patients received anti-cancer treatment during the study period, and all patients had an Eastern Cooperative Oncology Group (ECOG) performance status of 0-2 at inclusion. In addition, 12 cancer-free controls matched for sex, age, BMI, and cardiorespiratory fitness were included. Exclusion criteria were concurrent malignancy requiring active treatment, treatment with glucocorticoids (dexamethasone or prednisolone), history of thrombosis, BMI >30 kg/m², thrombocyte count ≤100 × 10⁹/L, and diabetes. All participants were screened for diabetes prior to inclusion and exhibited normal HbA1c levels.

At inclusion, anthropometric measurements (height and body weight) were obtained, BMI was calculated, body composition was assessed by dual-energy X-ray absorptiometry (DXA), and cardiorespiratory fitness was evaluated with a submaximal cardiopulmonary exercise test. Habitual physical activity monitoring was initiated, and fasting blood samples were collected, and analyzed at Department of Clinical Biochemistry, Rigshospitalet, Copenhagen, Denmark. Participant characteristics are found in Table S1.

#### Body composition

Body composition was assessed by DXA before and after the training intervention. All scans were conducted in the morning in fasting condition.

#### Submaximal cardiopulmonary exercise test

Cardiorespiratory fitness was assessed using the submaximal Ekblom-Bak test (26). The test was performed on a cycle ergometer (Lode Corival CPET) with continuous heart rate (HR) monitoring using a chest-worn HR sensor (Polar H10, Polar Electro, Finland). Participants cycled for four minutes at 32 W followed by four minutes at an individually adjusted workload corresponding to a HR of 110-140 beats per minute. Pedaling frequency was maintained at 60 revolutions per minute throughout the test. Estimation of VO_2_-max was calculated from the EB-test_2016_ sex-specific regression model (26).

#### Physical activity level

Habitual physical activity, defined as steps/day, was assessed using a thigh-mounted triaxial accelerometer (SENS motion®, SENS Innovation ApS). The sensor was attached to the lateral aspect of the non-trained thigh, approximately 10 cm above the knee, and worn for a minimum of seven consecutive days during the intervention. Participants were instructed to maintain their habitual physical activity level throughout the monitoring period.

#### Training intervention

Prior to the intervention, maximal endurance and strength tests were conducted to determine peak workload (PWL) and 10-repetition maximum (10RM), respectively. PWL was assessed during an incremental one-legged knee-extensor exercise test initiated at 10 W, with workload increased by 5 W every 5 min. Ratings of perceived exertion were assessed throughout the test using the Borg scale, and the test was continued until the participant was no longer able to maintain the required contraction frequency of one contraction per second.

To determine 10RM, participants performed six sets of one-legged knee extensions separated by two minutes of rest. The first five sets served as progressive warm-up sets, while the workload of the final set was estimated using ratings of perceived exertion (Borg scale) obtained during the preceding sets. Participants performed the final set to volitional fatigue, and 10RM was estimated from the load lifted and the number of completed repetitions using a modified version of the Brzycki equation (27) : 10RM = W * 0.7498 / (1.0278 – 0.0278 * r), where W is the load (kg) and r is the number of repetitions completed.

Participants subsequently completed a five-week supervised exercise intervention consisting of three weekly sessions of combined endurance and resistance one-legged knee-extensor exercise. Endurance training was performed on a modified Krogh bicycle ergometer and consisted of 40 minutes of dynamic knee-extensor exercise at an intensity corresponding to 60-120% of pre-intervention PWL. Resistance training consisted of six sets of ten knee extensions performed at 50-125% of pre-intervention 10RM, separated by two minutes of rest. All exercise sessions were supervised, and training intensity was adjusted throughout the intervention based on workload and ratings of perceived exertion assessed using the Borg scale. Following each session, participants received 125 mL of a protein supplement (Arla Protino® Plus; 12.5 g protein).

#### Hyperinsulinemic-euglycemic clamp

Participants underwent a metabolic test day 24 hours after the final exercise session to assess whole-body and leg-specific insulin sensitivity. During the 24 hours prior to the metabolic test day, participants followed a standardized diet providing 20E%, 44E%, and 36E% of energy from protein, carbohydrate, and fat, respectively, with total energy intake individually adjusted based on participant characteristics. Participants arrived in the morning following an overnight fast and remained fasted throughout the experimental procedures.

Vascular access was established by placement of catheters in an antecubital vein for infusion of insulin and glucose, in one femoral artery for arterial blood sampling, and bilaterally in the femoral veins for venous blood sampling. Baseline blood samples were obtained prior to insulin stimulation, and femoral artery blood flow was assessed bilaterally using panoramic ultrasound (NextGen LOGIQ™ e Ultrasound System). Indirect calorimetry was performed to assess basal metabolic rate and substrate utilization. Skeletal muscle biopsies were subsequently collected before initiation of a 2-hour hyperinsulinemic-euglycemic clamp.

The clamp was performed using a primed continuous insulin infusion (9 mU/kg bolus followed by 1.42 mU/kg/min), while glucose infusion rates were continuously adjusted to maintain euglycemia at fasting glucose concentrations. Throughout the clamp, arterial and femoral venous blood samples were obtained at 20. minute intervals, and concurrent blood flow measurements in the femoral arteries were performed in both legs to quantify leg-specific glucose uptake. Following 90-minutes of insulin stimulation, indirect calorimetry was assessed again and after 2-hour insulin stimulation, muscle biopsies were repeated. Participants were subsequently provided with a meal and remained under observation until blood glucose concentrations had returned to stable levels.

#### Indirect calorimetry

Participants were placed in the supine position in a hospital bed, where they rested for at least 30 minutes. Oxygen uptake and carbon dioxide production were measured with a Metalyzer 3B (Cortex Biophysik GmbH, Leipzig, Germany), which was calibrated before each test day. Participants wore a face mask, and expired air was collected for 10 minutes; the first 5 minutes were discarded, and the average of the last 5 minutes was used.

#### Arterial blood flow

Arterial blood flow was measured using doppler ultrasound on a NextGen LOGIQ™ e (GE healthcare, Chicago, IL, USA) scanner using a linear probe (12L-RS Probe, GE healthcare, Chicago, IL, USA). Arterial blood flow was measured for at least 15 seconds just prior to the blood samples.

#### Calculating leg glucose uptake

To calculate glucose uptake (LGU) across each leg, the arterial-venous glucose difference was multiplied with the arterial blood flow (Fick’s Principle) from the measurements obtained every 20. minute. LGU was then divided by lean leg mass obtained from the DXA-scan obtained in the morning on the experimental day. Insulin-stimulated glucose uptake was expressed as area under the curve during the last 40 minutes of the hyperinsulinemic euglycemic clamp (AUC_40 min_).

#### Blood sample handling

Blood samples were immediately centrifuged at 3000 g for 10 minutes, and plasma was transferred into Sarstedt tubes and stored at -70°C until further processing.

#### Muscle biopsies

Skeletal muscle biopsies were obtained from the vastus lateralis under local anesthesia (10 mg/mL lidocaine) from both legs. The biopsy site was located approximately one-third of the distance between the patella and the inguinal fold. Following a small skin incision, a 6-mm Bergström needle was inserted through the fascia into the muscle, and tissue samples were collected using manual suction.

#### Muscle glycogen concentration

Muscle glycogen concentration was determined as glycosyl units after acid hydrolysis by a fluorometric method (28).

#### Muscle sample preparation

Muscle samples (≤25 mg) were pulverized on dry ice, weighed, and transferred to 2 mL Eppendorf tubes. Tissue was homogenized in ice-cold MG buffer (10% glycerol, 20 mM Na-pyrophosphate, 150 mM NaCl, 50 mM HEPES pH 7.5, 1% NP-40, 20 mM β-glycerophosphate, 10 mM NaF, 2 mM PMSF, 1 mM EDTA pH 8.0, 1 mM EGTA pH 8.0, 10 μg/mL aprotinin, 10 μg/mL leupeptin, 2 mM Na₃VO₄, 3 mM benzamidine and dD H2O) using a TissueLyser (Qiagen, USA) with stainless steel beads for 60 seconds at 30 Hz. Homogenates were incubated under end-over-end rotation for 30 min at 4°C and subsequently centrifuged at 18,000 × g for 20 min at 4°C. The resulting supernatant was collected and transferred to fresh tubes. Samples were kept on ice throughout the procedure and stored at −70°C until further analysis. Total protein concentration was determined using Pierce bicinchoninic acid (BCA) assay (Thermo Fisher Scientific).

#### Immunoblot procedure

Immunoblotting was performed on muscle tissue to determine protein abundance and phosphorylation of selected proteins involved in insulin signaling and glucose metabolism.

#### Laemmli samples

Muscle lysates were diluted to a final protein concentration of 2 μg·μL⁻¹, determined by bicinchoninic acid (BCA) assay, using 6× Laemmli sample buffer (0.5 M Tris base, pH 6.8, 20% glycerol, 11% SDS, 0.05% bromophenol blue, and 225 mM DTT).

#### SDS-PAGE

Equal amounts of protein (10-20 μg) were separated on 7, 10, 12, or 4-15% Mini-PROTEAN TGX polyacrylamide gels (Bio-Rad). A standard curve consisting of increasing protein was included on each gel to verify linearity and loading accuracy. Electrophoresis was performed at 150 V for 45-60 minutes. Following separation, proteins of interest were transferred to polyvinylidene difluoride (PVDF) membranes (Immobilon, Millipore) using a semi-dry transfer system (Trans-Blot Turbo, Bio-Rad) at 14 V and 2.5 A for 45 minutes, or 10 V and 2.5 A for 80 minutes depending on the molecular weight of the target protein. Membranes were blocked in 2% skim milk prepared in Tris-buffered saline containing 0.1% Tween-20 (TBST pH 7.4) and incubated overnight at 4°C with primary antibodies (Table S3). Following washing in TBST, membranes were incubated with horseradish peroxidase-conjugated secondary antibodies for 45 min at room temperature and washed prior to detection.

#### Imaging and quantification

Protein bands were visualized by enhanced chemiluminescence (Immobilon Forte Western HRP Substrate, Millipore) and imaged using a ChemiDoc MP Imaging System (Bio-Rad). Band intensities were quantified using Image Lab software (version 6.1) and corrected for background signal. A standard sample was included on each membrane to verify transfer consistency.

#### HOMA index:

The homeostatic model assessment of insulin resistance (HOMA) was calculated as fasting insulin (µU/mL) × fasting glucose (mmol/L) / 22.5.

#### Plasma insulin, c-peptide, and fatty acid concentrations

Insulin concentrations were measured using human insulin enzyme-linked immunosorbent assay (ELISA) (Mercodia, Uppsala, Sweden; cat. no. 10-1113-01). C-peptide concentrations were measured in plasma samples using the Roche Cobas 6000 platform. Fatty acids were measured enzymatically using the NEFA-HR(2) kit (Fujifilm Wako Chemicals, Neuss, Germany; R1: 434-91795, R2: 436-91995).

#### Statistical analysis

Statistical analyses were performed using R (version 4.6.1) with the packages lme4 (1.1-35), lmerTest (3.1-3) and emmeans (1.10.0). Data are presented as individual values with mean ± SD. All analyses were adjusted for sex. Outcomes with one value per participant were compared between patients and controls using linear models with group and sex as fixed effects. Variables involving repeated measurements, including the effects of training (trained vs. non-trained leg), insulin stimulation (basal vs. insulin-stimulated conditions), group (patients vs. controls), and their interactions, were analyzed using linear mixed-effects models with group, training or insulin, their interaction and sex as fixed effects and participant as a random intercept (paired with-in subject design). Participants with a missing value in one condition were retained in the mixed models. When the interaction had p<0.05 (p=0.053 in Fig. 2B), Sidak-adjusted post hoc comparisons were performed. The composite protein score was calculated for each participant and leg as the mean of the six proteins shown (GLUT4, HKII, Akt2, GSK3β, TBC1D4 and AMPKα2), each expressed relative to the non-trained leg of the controls. Normality of model residuals was assessed using the Shapiro–Wilk test. Associations between variables were assessed using partial Pearson correlation coefficients adjusted for sex, calculated by correlating the residuals of both variables after regression on sex. Spearman's rank correlation, adjusted for sex in the same way, was used for CRP because most values were at the detection limit. Correlation analyses were exploratory and were not adjusted for multiple comparisons. Statistical significance was accepted at p<0.05.
